# Detecting Goals of Care Conversations in Clinical Notes with Active Learning

**DOI:** 10.1101/2024.01.03.24300801

**Authors:** Davy Weissenbacher, Katherine Courtright, Siddharth Rawal, Andrew Crane-Droesch, Karen O’Connor, Nicholas Kuhl, Corinne Merlino, Anessa Foxwell, Lindsay Haines, Joseph Puhl, Graciela Gonzalez-Hernandez

## Abstract

**Objective:** Goals Of Care (GOC) discussions are an increasingly used quality metric in serious illness care and research. Wide variation in documentation practices within the Electronic Health Record (EHR) presents challenges for reliable measurement of GOC discussions. Novel natural language processing approaches are needed to capture GOC discussions documented in real-world samples of seriously ill hospitalized patients’ EHR notes, a corpus with a very low event prevalence.

**Methods:** To automatically detect utterances documenting GOC discussions outside of dedicated GOC note types, we proposed an ensemble of classifiers aggregating the predictions of rule-based, feature-based, and three transformers-based classifiers. We trained our classifier on 600 manually annotated EHR notes among patients with serious illnesses. Our corpus exhibited an extremely imbalanced ratio between utterances discussing GOC and utterances that do not. This ratio challenges standard supervision methods to train a classifier. Therefore, we trained our classifier with active learning.

**Results:** Using active learning, we reduced the annotation cost to fine-tune our ensemble by 70% while improving its performance in our test set of 176 EHR notes, with 0.557 F1-score for utterance classification and 0.629 for note classification.

**Conclusion:** When classifying notes, with a true positive rate of 72% (13/18) and false positive rate of 8% (13/158), our performance may be sufficient for deploying our classifier in the EHR to facilitate point-of-care access to GOC conversations documented outside of dedicated notes types, without overburdening clinicians with false positives. Improvements are needed before using it to enrich trial populations or as an outcome measure.

## 1. Significance and Background

Discussing and documenting patients’ goals of care in serious illness is guideline-recommended and associated with improved patient - and family - reported outcomes and end-of-life care Wright et al. (2008); Bernacki et al. (2014); Detering et al. (2010). GOC discussions have also been identified as a key quality metric in serious illness care of Medicine (2015) and outcome measure for studies of communication and palliative care interventions Sudore et al. (2018). However, GOC discussions remain infrequent among many seriously ill populations, and even when it does occur, it is often in the final weeks or days of life thus limiting its benefit. Quality improvement and research efforts to improve this status quo have been hampered by the lack of an accurate, reliable, and scalable method for identifying GOC discussions documented infrequently and inconsistently by clinicians in the electronic health record (EHR).

Some hospitals include in their EHR a dedicated note type or area in the chart for documenting GOC discussions. For example, in our study health system’s inpatient EHR, there is an Advance Care Planning (ACP) note type that clinicians are encouraged to use for documenting GOC discussions. This is intended to facilitate standardization and accessibility of these important conversations at the point-of-care, and promote continuity of care across different healthcare settings and clinicians. However, clinicians commonly alternatively document GOC discussions within other notes that quickly become “buried” among the vast data contained within a patient’s inpatient chart Lee et al. (2021). This has both practical and scientific implications for patient care and the accuracy and reliability of documented GOC as a key patient-centered outcome, respectively.

In this study, relying on Natural Language Processing (NLP) methods, we present an automatic classifier to retrieve GOC discussions in the EHR that may facilitate both clinicians’ point-of-care access to their patients’ documented goals (or lack thereof) and arduous data extraction processes for research. Recent studies have employed various NLP methods to identify GOC discussions in the EHR Chan et al. (2019); Lee et al. (2021); Davoudi et al. (2022) in different care settings and seriously ill populations. All of the studies reported good overall model performance in internal validation cohorts and a substantial reduction in human annotation time for EHR review. Collectively, these studies suggest that NLP offers a promising and powerful approach to overcome EHR documentation challenges. However, the prior NLP methods have been developed on datasets that were purposively enriched for GOC discussions (e.g., patient selection, note type selection, definition of GOC discussions, and thus do not represent real world EHR data in which documentation of GOC discussions is present much less commonly. Here, our goal is to automate the retrieval of GOC discussions documented *anywhere* in EHR clinical notes among a general hospital population with serious illness.

## 2. Materials and Methods

We expressed the task of retrieving GOC discussions documented in EHR as a binary classification task. We aim to train a classifier to retrieve all utterances in clinical notes that are documenting a GOC conversation. To train and evaluate our classifier, we collected 776 inpatient clinical notes, 400 Advance Care Planning notes, and 376 notes of different types, hereafter called general notes, from the EHR of patients with an advanced serious illness (solid malignancy, chronic pulmonary disease, chronic kidney disease, heart failure, chronic liver disease, and neurodegenerative disease) who were admitted to one of five hospitals in the University of Pennsylvania Health System between April 1, 2017 and December 31, 2019. We sampled the notes with a stratified strategy to ensure a representative distribution across severity of illness, Courtright et al. (2019) race, and note type. Two trained annotators (NK, medical resident and CM, research coordinator) independently labeled each utterance of the notes as being a part of a GOC conversation or not. A palliative care specialist (KC or AF) adjudicated disagreements. The annotation guidelines are available here. We found moderate agreement between the two annotators on both types of notes (with a 0.685 Cohen’s Kappa score McHugh (2012) on the ACP notes and 0.659 on the general notes). As anticipated, there was a higher volume of GOC conversations in ACP notes compared to general notes. After adjudication, we found in the ACP notes that 1,761 utterances were labeled as belonging to a GOC conversation (the positive examples) and 14,489 utterances labeled as not belonging to a GOC conversation (the negative examples), leading to an imbalance ratio of 9/1; in the general notes we also found GOC conversations with 109 utterances labeled positive, but most sections of the notes (24,174 utterances) were not part of a GOC discussion, resulting in a much higher imbalance ratio of 155/1. This extreme imbalance challenges supervised learning methods for automatic classification as the negative examples, utterances not part of a GOC discussion, dominate.

To automate the annotation, we trained our classifiers with supervision, as it is currently the learning method that achieves the best performance on classification tasks Jimenez Gutierrez et al. (2022). However, since our dataset is extremely imbalanced, we could not follow the general practices to train classifiers. These practices are designed for balanced datasets. Imbalanced datasets present well-known challenges for training classifiers Fernández et al. (2018). First, the cost of annotation is higher for imbalanced datasets, because most utterances annotated are negative examples which are providing no information about the linguistic patterns used by healthcare providers when documenting GOC conversations in the notes. Our medical experts would spend most of their efforts labeling obvious negative examples, which would be easily discriminated by a classifier. Second, general learning algorithms are designed to learn from balanced datasets and tend to overgeneralize when applied to imbalanced datasets. That is, to minimize their loss during training, they assume that the few positive examples seen in the training set are noise and classify them as negative examples, resulting in poor performance when the detection of the positive examples is evaluated.

As an alternative to passive learning, the general training practice, we trained our classifier with active learning and showed its benefits when the datasets are extremely imbalanced. When applied to an imbalanced dataset, active learning is an efficient heuristic to re-balance the distribution between positive/negative examples and improve the learning process of a classifier. We compare passive and active learning in Figure 1. During our experiments, we evaluated both approaches. We created four datasets to train and evaluate our classifiers. First, we created a *seed set* with all utterances of the 400 Advance Care Planning notes dense with GOC conversations. From the 316 general notes, we randomly selected 200 notes and split the utterances, hiding from the classifiers the labels of 80% of the notes (the *unlabeled set*) and reserving the remaining 20% as the *validation set*. We set aside all utterances of the 176 remaining general notes as the *test set*. In the first series of experiments, we trained our classifiers with passive learning, the default method. We trained our classifier on the *seed set*, using the *validation set* for internal control, and evaluated it on the *test set*. In a second series of experiments, we trained our classifier with active learning; we started to train our classifier on the *seed set*, but we continued in an iterative and interactive process to train the classifier on a small set of examples in the *unlabeled set* that were hard to predict (uncertainty sampling Settles (2012)). Usually, those examples are the examples closest to the decision boundary as shown in Figure 1. At each iteration, we then retrained our classifier on the examples of the *seed set* plus the hard examples for which we released the labels. We stopped this selection process after five consecutive iterations if the classifier did not improve its performance on the validation set. We evaluated on the *test set* the model of the classifier computed during the best iteration on the *validation set*. We ran a last series of experiments to control the performance of the classifier when it was trained with passive learning on all examples available, i.e. all examples in the *seed set* and in the *unlabeled set*.

**Figure 1:**
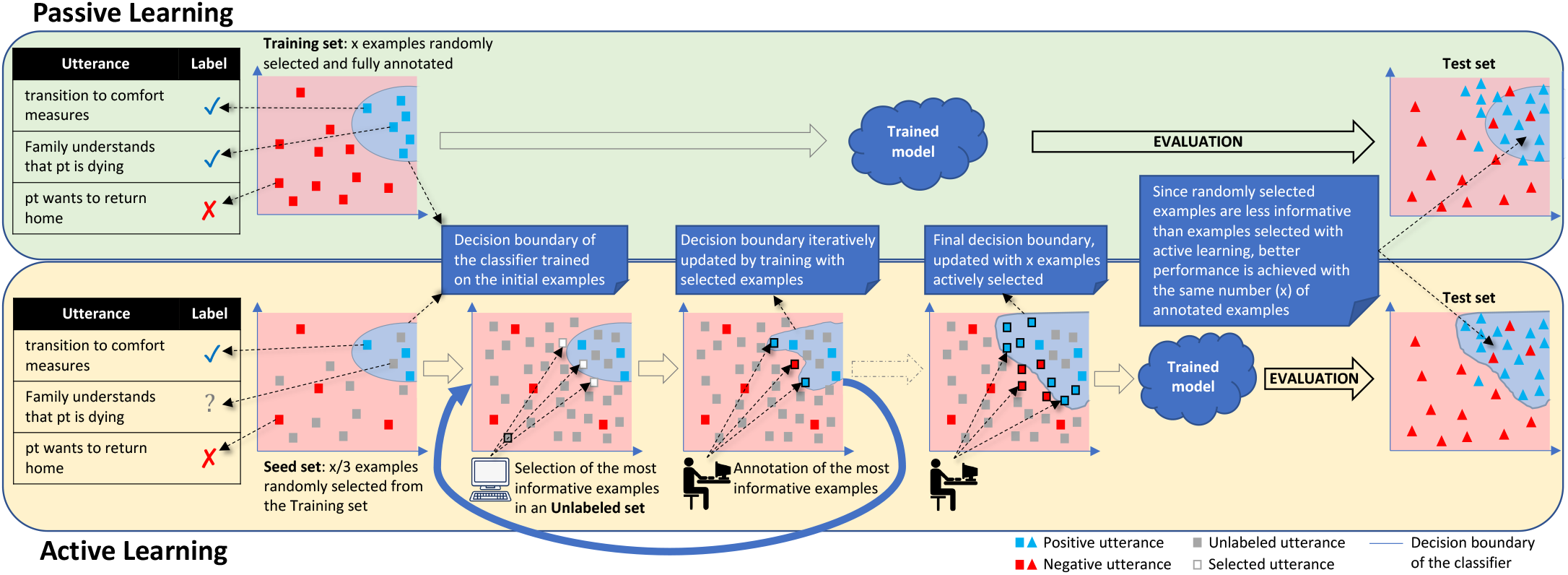
Passive *vs* Active learning. In passive learning, a classifier is trained on a sample of utterances randomly selected and annotated. In active learning, a classifier is trained through iterative interactions with an annotator. During an iteration, the annotator only annotates the utterances close to the decision boundary of the classifier defined for the iteration; the classifier is then retrained to update its boundary to include the newly annotated positive utterances. All utterances in the blue zone delimited by a decision boundary are predicted as positive by the classifier, utterances in the red zone are predicted negative. (Note: we reduced the size of the vectors to two dimensions for visualization)

We used an ensemble of five classifiers to detect the utterances documenting a GOC conversation: a regular expressions classifier, a logistic regression classifier working with filtered uni/bi-grams, three transformer classifiers working with contextual word embeddings (BERT-base cased, BioClinical BERT, and RoBERTa). We evaluated three methods to aggregate their predictions. With the simplest method, the average prediction, we averaged the probabilities measuring the certainty of each classifier when predicting the positive class. When the average was above 0.5, the ensemble predicted the positive class, the negative class otherwise. Since some classifiers achieved higher performance than others on the validation set, we found it beneficial to assign more weight to the most reliable classifiers when averaging the predictions of all classifiers in our second aggregating method, weighted average prediction. We weighted the certainty of the prediction of each base classifier by the F1-scores they achieved on the validation set. For our last method, we trained a decision tree to choose the best predictions within the context of each example. We trained the decision tree to predict the labels of all training examples given the features describing the predictions of each base classifier: the unigrams, the bigrams of the utterances, and the predictions of the base classifiers represented by nominal categories. We binned the certainty of each base classifier into a very-unlikely bucket when the certainty *x* was in the interval 0 <= *x* < 0.25, an unlikely bucket in the interval 0.25 <= *x* < 0.5, likely in 0.5 <= *x* < 0.75, and very-likely in 0.75 <= *x* <= 1. We also evaluated the performance of the base classifiers individually on our task.

## 3. Results

We ranked our classifiers according to their ability to retrieve the individual utterances belonging to a GOC conversation as well as to their ability to retrieve clinical notes recording a GOC conversation. We measure their performance with a metric well accepted for imbalanced datasets Fernández et al. (2018), the F1-score computed on the positive class, the utterances/notes labeled as belonging to/reporting a GOC conversation. We run each experiment three times to take into account the stochastic nature of the training process of neural networks and reported the mean of their scores in Table 1.

**Table 1:** Classification of utterances and clinical notes discussing Goal of Care. In the *default* setting, classifiers were only trained on Advance Care Planning notes; in the *active learning* setting, classifiers were trained on Advance Care Planning notes and the most informative examples queried from the general notes; in the *all examples* setting, classifiers were trained on all examples available from Advance Care Planning and the general notes. For the non-deterministic classifiers, we reported the means of their scores achieved during three runs.

| Training | Classifier | utterance classification |  |  | note classification |  |  |
| --- | --- | --- | --- | --- | --- | --- | --- |
|  |  | P | R | F1 | P | R | F1 |
| Default | RegEx | 0.2048 | **0.7907** | 0.3254 | 0.2698 | **0.9444** | 0.4198 |
|  | Logistic Regression | 0.102 | 0.3488 | 0.1579 | 0.2222 | 0.6667 | 0.3333 |
|  | BERT-base | 0.091 | 0.6 | 0.153 | 0.2115 | 0.9 | 0.34228 |
|  | BioClinical BERT | 0.0734 | 0.6884 | 0.1324 | 0.2115 | 0.9 | 0.34228 |
|  | RoBERTa-base | 0.0765 | 0.6977 | 0.1379 | 0.191 | **0.9444** | 0.3178 |
|  | Ensemble-average prediction | 0.1788 | 0.6124 | 0.2756 | 0.2798 | 0.8148 | 0.4165 |
|  | Ensemble-weighted average prediction | 0.3595 | 0.6202 | 0.4518 | 0.3888 | 0.6667 | 0.4853 |
|  | Ensemble-decision tree | 0.0719 | 0.6279 | 0.1291 | 0.2189 | 0.8889 | 0.351 |
| Active Learning | Logistic Regression | **0.6552** | 0.4419 | 0.5278 | **0.5714** | 0.4444 | 0.5 |
|  | Ensemble-average prediction | 0.4940 | 0.6202 | 0.5499 | 0.5357 | 0.7037 | 0.6079 |
|  | Ensemble-weighted average prediction | 0.4852 | 0.6434 | **0.5532** | 0.5488 | 0.7592 | **0.6359** |
|  | Ensemble-decision tree | 0.4474 | 0.5931 | 0.51 | 0.54 | 0.75 | 0.6279 |
| All examples | Logistic Regression | 0.6176 | 0.4884 | 0.5455 | 0.5294 | 0.5 | 0.5143 |
|  | Ensemble-average prediction | 0.4554 | 0.6202 | 0.5247 | 0.5521 | 0.7037 | 0.6182 |
|  | Ensemble-weighted average prediction | 0.4423 | 0.6124 | 0.513 | 0.5224 | 0.6852 | 0.5921 |
|  | Ensemble-decision tree | 0.385 | 0.5814 | 0.458 | 0.5209 | 0.75 | 0.6139 |

What stands out from the table 1 is that the identification of GOC conversations in general clinical notes is still a challenging problem. Our best classifier, an ensemble of 5 base classifiers aggregating their predictions with a weighted average and trained with active learning, achieved moderate F1 scores of 0.557 for the utterance classification and 0.629 for the notes classification. However, while moderate, we judge the performance of our classifier high enough to be deployed in production. Looking at the details of the confusion matrix of the best run of our ensemble when classifying clinical notes, we can see that it classified correctly the large majority of the notes which do not discuss a GOC in the test set, with 145 negative notes correctly identified. Out of the 18 notes discussing a GOC the ensemble correctly identified 13 notes, missing only 5 notes, therefore retrieving 72% of the notes of interest. It incorrectly identified 13 notes as mentioning a GOC where they did not, i.e. 8% (13/158). We submit that this noise is small enough for a clinician to quickly review the notes detected by the classifier manually to facilitate patient care in real-time. In comparison, the simplest system, the RegEx classifier, retrieved all notes but one. However, it was at the cost of more errors, with 43 notes not discussing a GOC and incorrectly flagged by the RegEx classifier as doing so, requiring more time from the clinician to review these notes. In future work, if we give a higher priority to retrieving all notes discussing a GOC, we could design a dedicated interface to facilitate the curation of the notes retrieved by the RegEx by presenting the utterances ranked by the confidence of the ensemble.

It is apparent from Table 1 that we improved the F1-scores of our classifiers by training them with active learning. When we trained our classifiers with the default setting, we achieved good Recall with scores above 0.60, but very low Precision with scores under 0.35. To understand this low Precision, we analyzed their False Positives (FPs). Our classifiers were too sensitive to phrases frequently used in GOC conversations, such as *want, continue, for now, were discussed, etc*.. In the ACP notes that we used for training our classifiers, these phrases were reliable indicators since most comments in the notes are part of a GOC discussion. However, in general notes these words are more often used in other contexts and cause the classifiers to erroneously recognize parts of GOC discussions when the classifiers are applied without fine-tuning their training on those notes. The FPs detected by our classifiers were utterances recording routine treatment and discharge plans, and minor care preferences unrelated to GOC. When we trained the classifiers with active learning those utterances were the first utterances queried by the classifiers allowing them to improve their Precision from 20 points during the first 3 iterations, before plateauing as shown by Figure 2.

**Figure 2:**
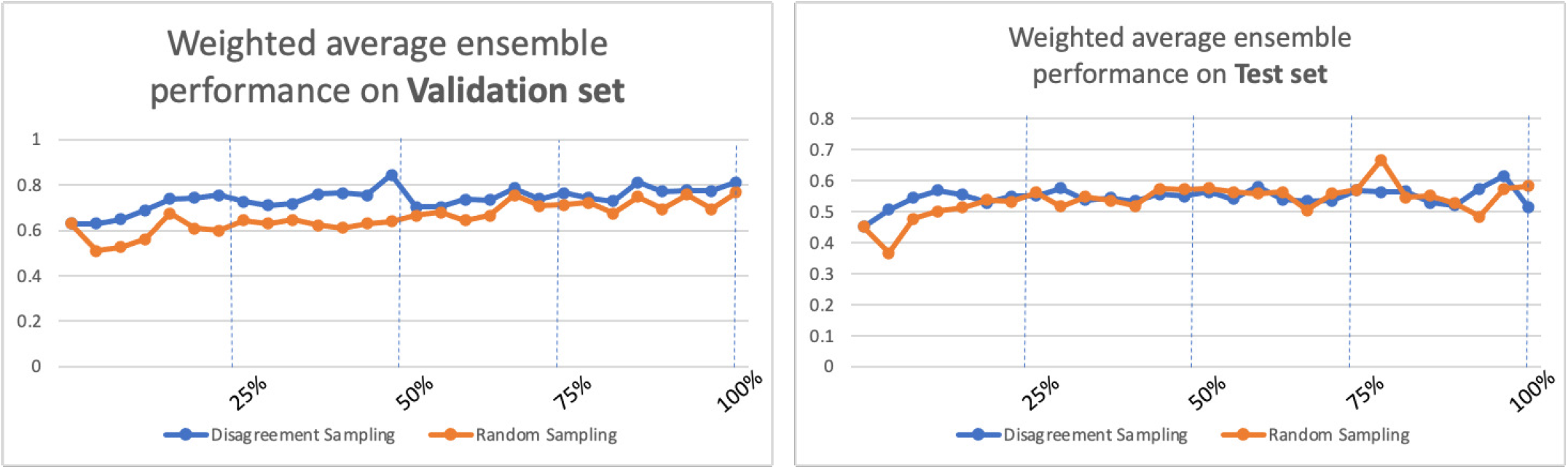
Performances of the Weighted average ensemble with increasing numbers of training examples from the general clinical notes. The performances reported are the average F1-scores achieved during 3 different runs by the ensemble using two different sample strategies during learning, disagreement and random samplings.

Interestingly, with the exception of the Logistic Regression classifier, we observed that when we trained our classifiers with all examples annotated, during the *all examples* setting, the classifiers did not achieve better performance than when we trained them only on the examples they selected through active learning. A possible explanation might be the effective change in the distribution of the examples done during the selection of the active learning. When we trained a classifier with active learning, the classifier only focuses on the examples of the ACP notes, which has a low imbalance ratio, and the few examples selected during the first iterations of the training by the algorithms in the *unlabeled set*. Since we stopped the training of the classifier when its performance on the *validation set* shows no improvement despite more annotated examples being provided for training, the imbalance ratio remains low. This early stopping criteria allows us to train the classifier on a more balanced dataset, avoiding the pitfall of training on a skewed dataset with the classifier only focusing on the negative examples to optimize its loss. Moreover, despite being few in number, since the examples selected in the *unlabeled set* are the most challenging for the classifier, they are the most informative for training. These examples are showing important linguistic patterns to improve the classification, leaving redundant or obvious negative examples out of the selection.

Finally, with active learning, we were also able to reduce significantly the annotation effort without compromising with the overall performance of our classifiers. When training the best ensemble with active learning during the three runs for its evaluation, we found that our early stopping criteria stopped the training around the 8^*th*^ iteration in average (σ = 4.1), resulting in 3,200 additional utterances to be annotated. That is, we only needed to annotate 30% (3,200/10,541) of the utterances from the general notes in the *unlabeled set* to guarantee an improvement of the ensemble of classifiers when classifying these notes. Note that we excluded the cost of annotation from the *seed set* when computing the reduction of the annotation effort, only considering the reduction of the annotation cost for the utterances from the *unlabeled set*. To test our hypothesis, that is, a classifier trained only on ACP notes will under-perform when retrieving the GOC conversations in general notes which are the notes of interest for this study, we fully annotated the ACP notes constituting our the *seed set* and did not use AL to reduce the annotation cost for this set.

## 4. Discussion

In this study, our objective was to automatically detect utterances documenting GOC discussions in general clinical notes. Our results show that a classifier trained on ACP notes *only* will under-perform when retrieving the GOC conversations in general notes because the other types of conversations in the general notes confuse the classifier. We successfully used active learning to select examples of these reports of GOC conversations to fine-tune our classifier, an ensemble of 5 base classifiers, and significantly improve its performance on general notes while keeping the number of these additional examples to be labeled by our annotators within an acceptable limit of 3,200 utterances (i.e. 30% of the utterances from the *unlabeled set*).

However, despite a noticeable improvement, the performance of our ensemble remains moderate. A main reason for this could lie in its structure. We designed it to predict the label of an utterance by only presenting the utterance as input to the base classifiers. We made this choice due to the current technical limitations of the neural networks which compose our ensemble. The transformer model on which rely three base classifiers of our ensemble only accepts sequences of a maximum length of 512 tokens. As a consequence, the ensemble only computes features that model the meaning of one utterance and ignores the surrounding utterances. This view is local and incomplete since GOC conversations are often reported within a paragraph. A solution could be to retrieve the paragraphs documenting a GOC discussion with a generative model like chat-GPT Ouyang et al. (2022). However, we left the evaluation of this approach as future work since large language models need to be fined-tuned on clinical notes to achieve competing performance Jimenez Gutierrez et al. (2022), a process that is still an economic and technical challenge Scao et al. (2022).

Lastly, the benefit of active learning was limited during our experiments. In our setting, we randomly sampled a small set of clinical notes and fully annotated these notes, the *unlabeled set*. We used these notes to create a unique pool of unlabeled examples by hiding the labels of the examples from the classifiers and only reveal their labels during the active training of the classifiers. By creating a pool with a fixed set of examples, we simplified the annotation process, reduced its cost, and reduced the computation time to improve the speed of the active learning iterations. We also ensured that all classifiers had access to the same pool of examples when comparing their individual performances. However, by doing so, we restricted the exploration of the classifiers. They were only able to query for the labels of the positive examples occurring in the pool. This is a strong limitation since our dataset is imbalanced, with only 51 positive examples and 10490 negative examples in our pool. We intend to repeat our experiments with the full dataset, 3 million unlabeled notes available at the Hospital of the University of Pennsylvania. We hypothesize that with more examples of reported conversations occurring in the dataset, our classifiers would be able to query for their labels and better discriminate the reports of GOC conversation from reports of other conversations, thus improving quickly their precision.

## 5. Related Work

The automatic detection of GOC conversations in clinical notes is a relatively new problem for the NLP community. In the last 5 years, researchers have released 11 systems that attempt to identify GOC conversations in clinical notes. The systems’ performance is compared to manual reviews of the charts and deployed on various cohorts to assess the frequencies of those conversations. The systems range from simple keyword matching approaches Lee et al. (2020); Brizzi et al. (2019) and hand-crafted, feature-based classifiers Uyeda et al. (2022); Lee et al. (2021), to advanced deep neural networks Davoudi et al. (2022); AlBashayreh (2022); Udelsman et al. (2019); Chan et al. (2019); Chien et al. (2019); Chien (2017). However, the performance of the systems cannot be compared to ours given that annotated datasets that can be used for development and evaluation are not shared. Authors that use hospital data are bound by privacy concerns, particularly with unstructured data, and those that use de-identified MIMIC III electronic records still requiring sharing fully annotated datasets. Further, variations in the experimental settings across these studies, particularly the definitions of what is considered a GOC conversation and the selection criteria for the cohort make it impossible to arrive at an “apples to apples” comparison.

A limitation of comparing NLP systems across studies is that no standard or consensus definition of what constitutes a GOC discussion exists. Thus, studies have used annotation guidelines derived by local experts, including the study reported herein. There are common themes across each guideline - care preferences, goals, and values expressed by patients and families - however, the implementation of this general definition varies greatly across studies due to differences in precise definitions. For example, one study labeled an isolated mention of code status Chan et al. (2019) as a positive example of a GOC discussion, and another positively labeled instances when a clinician gave a patient a blank advance directive document Lee et al. (2021). While these concepts may be part of a GOC discussion, in isolation of a discussion about patients goals and values many experts in the field do not consider them sufficient. Moreover, code status is captured as structured data in most EHRs negating the need for an NLP system. Using weak supervision, Davoudi et al. Davoudi et al. (2022) trained their classifiers to predict the predefined labels associated with the free-text content of comments extracted from a note template dedicated to a specific type of serious illness conversation. Because the authors did not evaluate their classifiers on unstructured general notes, its generalizability is quite limited Bernacki et al. (2015). From an NLP methods perspective, these prior annotation approaches result in a relatively high prevalence of positive examples in the corpora, which manifests in high performance achieved by the systems. This study used a stricter definition of a GOC discussion that required evidence in the clinical note of a discussion about goals or values, prognosis, and/or care preferences (code status documentation alone was not labeled as positive) between a clinician and patient or their surrogate. As anticipated and described in this report, this approach resulted in a very imbalanced dataset among the general clinical note, yet we argue that the trade off is high clinical applicability, patient-centeredness, and generalizability in real world EHR data.

The choice of the cohort to draw data from also impacts the balance of non-relevant to relevant discussions that need to be identified. For example, in AlBashayreh (2022); Lee et al. (2020); Chan et al. (2019) the authors collected the notes of critically ill patients, and in Davoudi et al. (2022) they focused on those with advanced cancer. Both of these populations are more likely to have had a GOC discussion compared to patients with other serious illnesses Beernaert et al. (2013). Therefore, the corpora exhibit a higher ratio of notes with a GOC discussion to notes without one. In Chan et al. (2019) the authors report a balance of 31% of notes with GOC conversations in their corpus, and in AlBashayreh (2022), after filtering the sentences with precise keywords, the author reports a balance as high as 46% of sentences with a GOC conversation. In Lee et al. (2021) the authors collected notes of seriously ill patients but also completed their corpus with notes written by palliative care specialists to achieve better balance. In our study, aside from using ACP notes to transfer linguistic knowledge to our classifiers, we randomly selected notes from patients with myriad types of serious illnesses without imposing additional assumptions or heuristics to increase the number of notes reporting GOC discussions. Therefore, given the extreme imbalance of our test set, which is only composed of general notes, our corpus is challenging but representative of the real setting in which the system will be deployed: unseen notes of patients with a serious illness followed at UPHS. We also re-implemented some of the approaches proposed in the existing studies as our baseline systems, namely the regex, logistic regression, BERT, and Bio-Clinical BERT classifiers, to estimate their performance on our corpus and compare them to the performance of our best classifier (an ensemble of various classifiers).

## 6. Conclusion

In this study, we proposed a state-of-the-art ensemble of binary classifiers to detect clinical notes documenting goal-of-care (GOC) discussions for patients with a serious illness followed at the Hospital of the University of Pennsylvania. Despite moderate performance, with an F1 score of 0.629, we believe that our classifier can be deployed in production as it was able to retrieve 72% (13/18) of the notes manually labeled in our test set as having a GOC discussion, bringing up only 13 notes incorrectly identified as having a GOC, a number small enough to avoid alert fatigue. Our results also indicate that a nearly exhaustive search for GOC conversations is possible semi-automatically by presenting to an annotator the notes retrieved by keyword matching and ranked according to the confidence of our ensemble to facilitate the curation. We are currently correcting the labels predicted by our ensemble on clinical notes from the MIMIC III database. Since these notes are de-identified we will be able to release the corpus for reproducibility.

## Acknowledgment

The authors declare that there are no competing interests.

## Ethics approval

This study protocol was reviewed by the Institutional Review Board at the University of Pennsylvania and determined to meet the eligibility criteria for exemption (45 CFR 46) and a waiver of the HIPAA authorization requirement was granted (45 CFR 164).

## Data availability

The clinical notes annotated for this study are Protected Health Information and not publicly available at this point.

## Code availability

The code and the models trained during the study are available from the corresponding author, DW, on reasonable request.

## Author contributions

DW designed the experiments, implemented the active learning layer and the ensemble, computed the models, analyzed the results, and is the first author of the manuscript. KC secured funding, guided the overall study design (in collaboration with GG-H for the automatic language processing aspects), wrote the annotation guidelines, validated the annotations, analyzed the results and wrote the portions of the manuscript relevant to the overview and significance of the problem. SR implemented the regular expression classifier. AC-D contributed to the study design, collected and preprocessed the data, wrote the dataset description and provided feedback on the manuscript. KO revised the annotation guidelines and computed the inter-annotator agreement. NK, CM, AF, LH and JP annotated the data. GG-H provided guidance overall and input on the design of the Natural Language Processing approaches used by DW and edited the manuscript.

## References

AlBashayreh, A.E., 2022. Evaluating Patient-Centered Outcomes in Palliative Care and Advanced Illness Using Natural Language Processing and Big Data Analytics. Ph.D. thesis. University of Iowa.

Beernaert, K., Cohen, J., Deliens, L., Devroey, D., Vanthomme, K., Pardon, K., Van den Block, L., 2013. Referral to palliative care in copd and other chronic diseases: a population-based study. Respir Med. 107, 1731–1739.

Bernacki, R., Block, S., of Physicians High Value Care Task Force, A.C., 2014. Communication about serious illness care goals: a review and synthesis of best practices. JAMA Intern Med 174, 1994–2003. doi:10.1001/jamainternmed.2014.5271.

Bernacki, R., Hutchings, M., Vick, J., Smith, G., Paladino, J., Lipsitz, S., Gawande, A., Block, S., 2015. Development of the serious illness care program: a randomised controlled trial of a palliative care communication intervention. BMJ Open 5. doi:10.1136/bmjopen-2015-009032.

Brizzi, K., Zupanc, S.N., Udelsman, B.V., Tulsky, J.A., Wright, A.A., Poort, H., Lindvall, C., 2019. Natural language processing to assess palliative care and end-of-life process measures in patients with breast cancer with leptomeningeal disease. Am J Hosp Palliat Care 37, 371–376.

Chan, A., Chien, I., Moseley, E., Salman, S., Kaminer Bourland, S., Lamas, D., Walling, A., Tulsky, J., Lindvall, C., 2019. Deep learning algorithms to identify documentation of serious illness conversations during intensive care unit admissions. Palliative medicine 33.

Chien, I., 2017. Natural Language Processing for Precision Clinical Diagnostics and Treatment. Ph.D. thesis. Massachusetts Institute of Technology.

Chien, I., Shi, A., Chan, A., Lindvall, C., 2019. Identification of serious illness conversations in unstructured clinical notes using deep neural networks, in: Koch, F., Koster, A., Riaño, D., Montagna, S., Schumacher, M., ten Teije, A., Guttmann, C., Reichert, M., Bichindaritz, I., Herrero, P., Lenz, R., López, B., Marling, C., Martin, C., Montani, S., Wiratunga, N. (Eds.), Artificial Intelligence in Health, Springer International Publishing. pp. 199–212.

Courtright, K., Chivers, C., Becker, M., Regli, S., Pepper, L., Draugelis, M., O’Connor, N., 2019. Electronic health record mortality prediction model for targeted palliative care among hospitalized medical patients: a pilot quasiexperimental study. J GEN INTERN MED 34, 1841–1847. doi:10.1007/s11606-019-05169-2.

Davoudi, A., Tissot, H., Doucette, A., Gabriel, P.E., Parikh, R., Mowery, D.L., Miranda, S., 2022. Using natural language processing to classify serious illness communication with oncology patients, in: Proceedings of AMIA Annual Symposium, pp. 168–177.

Detering, K.M., Hancock, A.D., Reade, M.C., Silvester, W., 2010. The impact of advance care planning on end of life care in elderly patients: randomised controlled trial. BMJ 340. doi:10.1136/bmj.c1345.

Fernández, A.H., García, S.L., Galar, M., Prati, R.C., Krawczyk, B., Herrera, F., 2018. Learning from Imbalanced Data Sets. Springer International Publishing.

Jimenez Gutierrez, B., McNeal, N., Washington, C., Chen, Y., Li, L., Sun, H., Su, Y., 2022. Thinking about GPT-3 in-context learning for biomedical IE? think again, in: Findings of the Association for Computational Linguistics: EMNLP 2022, Association for Computational Linguistics. pp. 4497–4512.

Lee, K.C., Udelsman, B.V., Streid, J., Chang, D.C., Salim, A., Livingston, D.H., Lindvall, C., Cooper, Z., 2020. Natural language processing accurately measures adherence to best practice guidelines for palliative care in trauma. Journal of Pain and Symptom Management 59, 225–232.e2.

Lee, R.Y., Brumback, L.C., Lober, W.B., Sibley, J., Nielsen, E.L., Treece, P.D., Kross, E.K., Loggers, E.T., Fausto, J.A., Lindvall, C., Engelberg, R.A., Curtis, J.R., 2021. Identifying goals of care conversations in the electronic health record using natural language processing and machine learning. Journal of Pain and Symptom Management 61, 136–142.e2.

McHugh, M., 2012. Interrater reliability: the kappa statistic. Biochem Med (Zagreb) 22, 276–82.

of Medicine, I., 2015. Dying in America: Improving Quality and Honoring Individual Preferences Near the End of Life. Washington (DC): National Academies Press (US).

Ouyang, L., Wu, J., Jiang, X., Almeida, D., Wainwright, C., Mishkin, P., Zhang, C., Agarwal, S., Slama, K., Ray, A., Schulman, J., Hilton, J., Kelton, F., Miller, L., Simens, M., Askell, A., Welinder, P., Christiano, P.F., Leike, J., Lowe, R., 2022. Training language models to follow instructions with human feedback, in: Koyejo, S., Mohamed, S., Agarwal, A., Belgrave, D., Cho, K., Oh, A. (Eds.), Advances in Neural Information Processing Systems, Curran Associates, Inc. pp. 27730–27744.

Scao, T.L., Fan, A., Akiki, C., et al., 2022. BLOOM: A 176B-Parameter Open-Access Multilingual Language Model. Working paper or preprint.

Settles, B., 2012. Active Learning. Synthesis Lectures on Artificial Intelligence and Machine Learning. Morgan & Claypool Publisher.

Sudore, R., Heyland, D., Lum, H., Rietjens, J., Korfage, I., Ritchie, C., Hanson, L., Meier, D., Pantilat, S., Lorenz, K., Howard, M., Green, M., Simon, J., Feuz, M., You, J., 2018. Outcomes that define successful advance care planning: A delphi panel consensus. J Pain Symptom Manage 55, 245–255. doi:10.1016/j.jpainsymman.2017.08.025.

Udelsman, B.V., Moseley, E.T., Sudore, R.L., Keating, N.L., Lindvall, C., 2019. Deep natural language processing identifies variation in care preference documentation. Journal of Pain and Symptom Management 59, 1186–1194.e3.

Uyeda, A.M., Curtis, J.R., Engelberg, R.A., Brumback, L.C., Guo, Y., Sibley, J., Lober, W.B., Cohen, T., Torrence, J., Heywood, J., Paul, S.R., Kross, E.K., Lee, R.Y., 2022. Mixed-methods evaluation of three natural language processing modeling approaches for measuring documented goals-of-care discussions in the electronic health record. Journal of Pain and Symptom Management 63, e713.

Wright, A., Zhang, B., Ray, A., Mack, J., Trice, E., Balboni, T., Mitchell, S., Jackson, V., Block, S., Maciejewski, P., Prigerson, H., 2008. Associations between end-of-life discussions, patient mental health, medical care near death, and caregiver bereavement adjustment. Journal of the American Medical Association 300, 1665–1673. doi:10.1001/jama.300.14.1665.

